# Prevalence of TEM and SHV Genes in *Klebsiella pneumoniae* isolated from different Clinical specimens in Khartoum, Sudan

**DOI:** 10.1101/2023.11.02.23297846

**Authors:** Saad A. S Hussein, Mutassim A.M. Salih, Hind M.E Adlan, Abualgasim E. Abdalla

**Author notes:** First Authors: Mutassim A.M Salih Hend M.E Adlan. Second author: Dr. Abualgasim Elgaili Abdalla PhD.

## Abstract

**Objective:** The aim of this study is to determine the prevalence of ESBL producing *Klebsiella pneumoniae* isolated from different clinical specimens gathered from different clinics in Khartoum State.

**Settings and Design:** This is a descriptive cross-sectional study conducted in Khartoum State during a period from February to May 2018.

**Methods and Materials:** *Klebsiella pneumoniae* isolated from patients attending to health-care settings with different infections, which was re-identified by conventional methods and screened for ESBL production by modified Kirby-Bauer’s method. The TEM and SHV genes were detected by PCR.

**Results:** Out of 100 *K. pneumoniae* isolates, 85% (85/100) were ESBL producing strains. The highest prevalence of ESBL production was in blood 100% (6/6) as well as stool 100% (4/4) followed by sputum 91% (21/23), wound 85% (12/14) and urine 82%(40\49) samples, while the lowest was noted in CSF 50% (2/4). The isolated *K. pneumoniae* showed high resistance to Ceftazidime 85% followed by Cefotaxime and Cefpodoxime 80% for each and 73% for Aztreonam. Furthermore, we found that 88% of isolates were resistant to Colistin and 33% were resistant to Meropenem. TEM and SHV genes were detected in 9.4% (8/85) and 1.2% (1/85) of ESBL producing strains, respectively. However, SHV was also detected in 6.7% (1/15) of non-ESBL-producing *K. pneumoniae*.

**Conclusions:** A high prevalence of ESBL-producing *K. pneumoniae* was detected in this study Which is 85%. TEM and SHV genes were found in some strains of ESBL producing *K. pneumoniae*. The findings of this study alarm the physicians to take ESBL producing strains of *K. pneumoniae* under consideration before making treatment decisions.

## Introduction

*Klebsiella pneumoniae* causes a wide range of hospital acquired infections, including pneumonia, urinary tract infections, septicemia, meningitis and soft tissue infections (1, 2). It comprised roughly 3-7% of all nosocomial bacterial infections (3). *K. pneumoniae* infections used to be treated promptly till the emergence of Extended-spectrum beta-lactamases (ESBL), which are enzymes that can hydrolyze broad-spectrum beta-lactam antibiotics (4). Parentally administered cephalosporins (cefotaxime, ceftazidime, ceftriaxone, and others) and a monobactam (aztreonam) with oxyimino side chains, mainly represented by gram-negative bacteria such as *Klebsiella pneumoniae, Escherichia coli, Salmonella* spp., *Proteus* spp., and related organisms (4), this is considered the main mechanism by which *K. pneumoniae* can resist the action of plethora of antibiotics (5). In Sudan the prevalence of ESBL producing *K. pneumoniae* was reported in 53% of strains isolated from urine specimens (6). Likewise, study in Zambia reported that a prevalence of ESBL was 52% (7). ESBLs are encoded by TEM and SHV genes which are harboring in plasmids (8, 9). The TEM was first reported in *E. coli* isolated from a patient named Temoniera in Greece, while SHV is referring to sulGhydryl variable active site (8). The prevalence of both genes among *K. pneumoniae* has been reported globally (10, 11, 12, 13). SHV was found in 67.1% (49/73) and TEM gene was only found in 16.4% of *K. pneumoniae* isolated from 7 European countries (14). Moreover, in Iran the frequency of SHV and TEM among the ESBLs producing isolates was 14.4% and 20.6%, respectively (15). Likewise, study conducted in India documented 67.3% of isolated *K. pneumoniae* have both SHV and TEM genes (16). Nonetheless, a study conducted in Sudan investigating the presence of ESBL in urine samples and the genes responsible for it, they detected SHV gene in 17.6% (2/68) and TEM gene in 16.2% (11/68) (17). Nevertheless, the prevalence of ESBL-producing *K. pneumoniae* or genes encoding ESBL in Sudan remains elusive. Therefore, the present study aimed to investigate the prevalence of ESBL producing *K. pneumoniae* and genes encoding ESBL, including TEM and SHV in strains isolated from various clinical specimens in Khartoum State.

## Materials and Methods

### Specimen collection and Isolation

This study included 100 non-duplicate isolates of *K. pneumoniae* from various clinical samples which were obtained from different hospitals in Khartoum State, namely, Fedail’s Hospital, Royal-Care Hospital, Albraha Hospital, Sharg-Elneil Hospital and Medical Military Hospital during a period from February to May 2018. The isolated organisms were identified by using conventional methods (colony morphology, Gram stain, oxidase test, citrate utilization test, motility test, urease test, indole test and sugars fermentation tests.

### ESBL phenotype detection

This was studied by Kirby-Bauer disc diffusion method as per guidelines from Clinical and Laboratory Standards Institute (CLSI) (18). BrieGly, to detect ESBL producing bacterial strains, 30μg Ceftriaxone, 30μg Cefotaxime, 30μg Aztreonam, 10μg Ceftazidime and 10μg Cefpodoxime (Bioanalyse, Turkey) were placed on Mueller-Hinton plates that had been inoculated with a *K. pneumoniae* isolate at density equivalent to 0.5% MacFarland standard turbidity. Additionally, two antibiotics were incorporated such 10μg Meropenem and 50μg Colistin (HiMedia, India) as treatment options. Plates were incubated overnight at 37 °C. Interpretation of the diameters of zones of inhibition was as described in [Table 1].

**Table 1:** The interpretation of the diameters of zones according to CLSI chart.

| **Antimicrobials** | **Sensitive (equal or more than)** | **Resistant (equal or less than)** |
| --- | --- | --- |
| Aztreonam 30µg | 21 | 17 |
| Cefpodoxime 10µg | 21 | 17 |
| Cefotaxime 30µg | 26 | 22 |
| Ceftriaxone 30µg | 23 | 19 |
| Ceftazidime 10µg | 21 | 17 |
| Meropenem 10µg | 23 | 19 |
| Colistin 50µg | 15 | 15 |

### DNA extraction and PCR

Genomic DNA was extracted from *K. pneumoniae* by phenol chloroform method as described previously (19). The purified DNA was preserved in TB EDTA at −20 °C till processing (20). TEM gene in about 861bp was amplified by using specific primers as follows; forward primer 5’-CATTTCCGTGTCGCCCTTATTC-3’, reverse primer 5’-CGTTCATCCATAGTTGCCTGA-3’. Specific primers for amplification of 471bp of the SHV gene included; forward primer 5’-AGCCGCTTGAGCAAATTAAAC-3’, reverse primer 5’-ATCCCGCAGATAAATCACCAC - 3’. The PCR mixture was prepared by adding 1μl from each primer (forward and reverse), 3μl of sterile dH_2_O, 5μl DNA sample to the 2μl of MasterMix (1.5 mM MgCl2, 200 μM each dNTP and 2-5 u Taq DNA polymerase) (Solis BioDyne, Estonia). Next, PCR mixtures were placed on thermocycler, which was optimized as followed at 95 °C for 5mins, annealing temperature for TEM was set at 59°C and for, SHV at 56.7 °C for 30sec, extension at 72 °C for 1min, final extension at 72°C for 10min. The PCR products were examined by gel electrophoresis as mentioned previously (21). [Figure 2].

## Results

A total of 100 *K. pneumonia* strains were isolated from different clinical specimens, including urine (n=49), sputum (n=23), wound (n=14), Blood (n = 6), CSF (n = 4) and Stool samples (n = 4).

This study revealed that the organisms were highly resistant to Colistin 88% and Ceftazidime 85% followed by Cefotaxime and Cefpodoxime accounting (80%), 74% were resistant to Ceftriaxone and 73% were resistant to Aztreonam, a lower resistance was observed for Meropenem 33%. [Figure 1].

**Figure 1.**
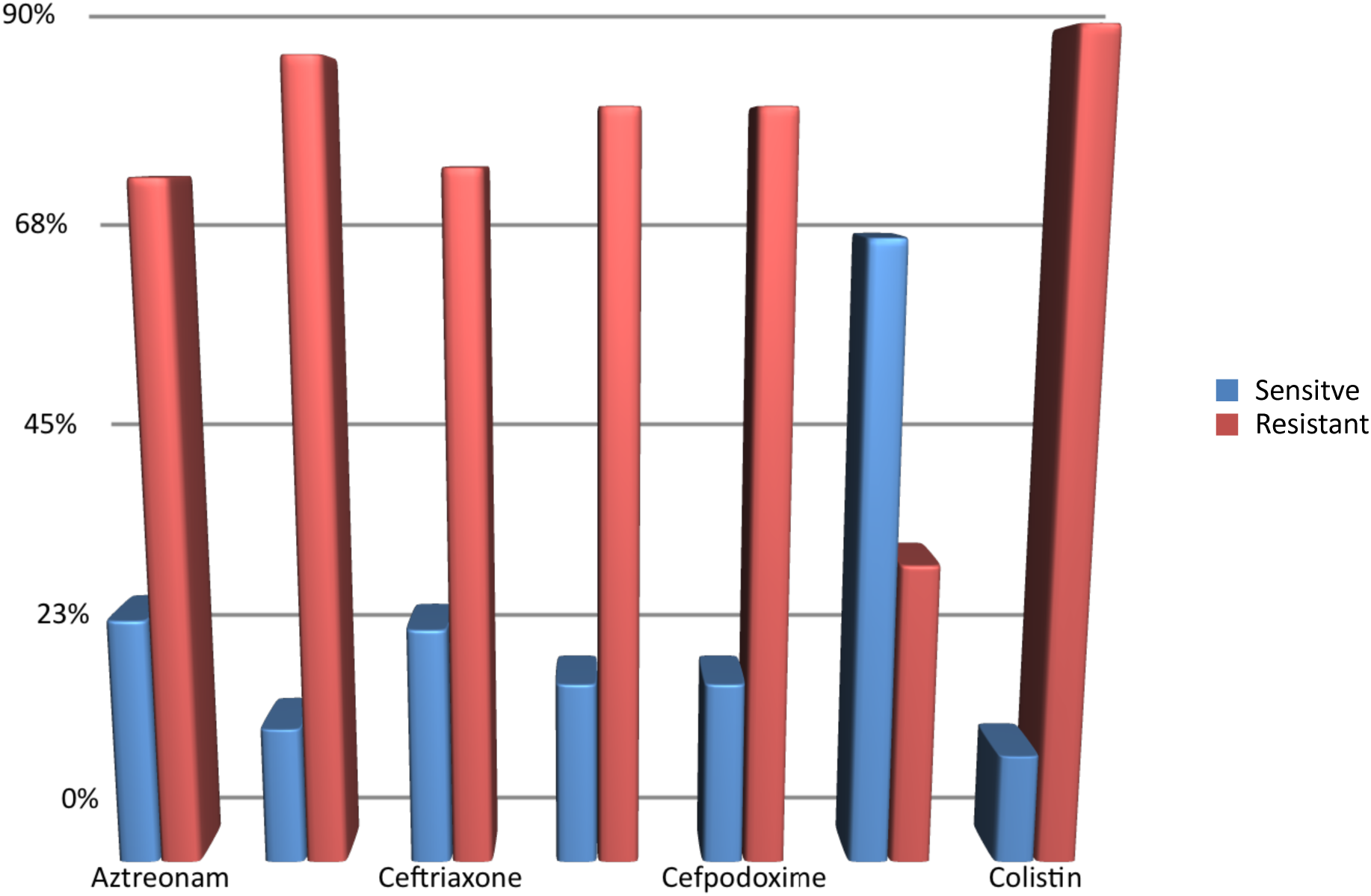
Antibiotic resistance patterns of *K. pneumoniae*.

**Figure 2:**
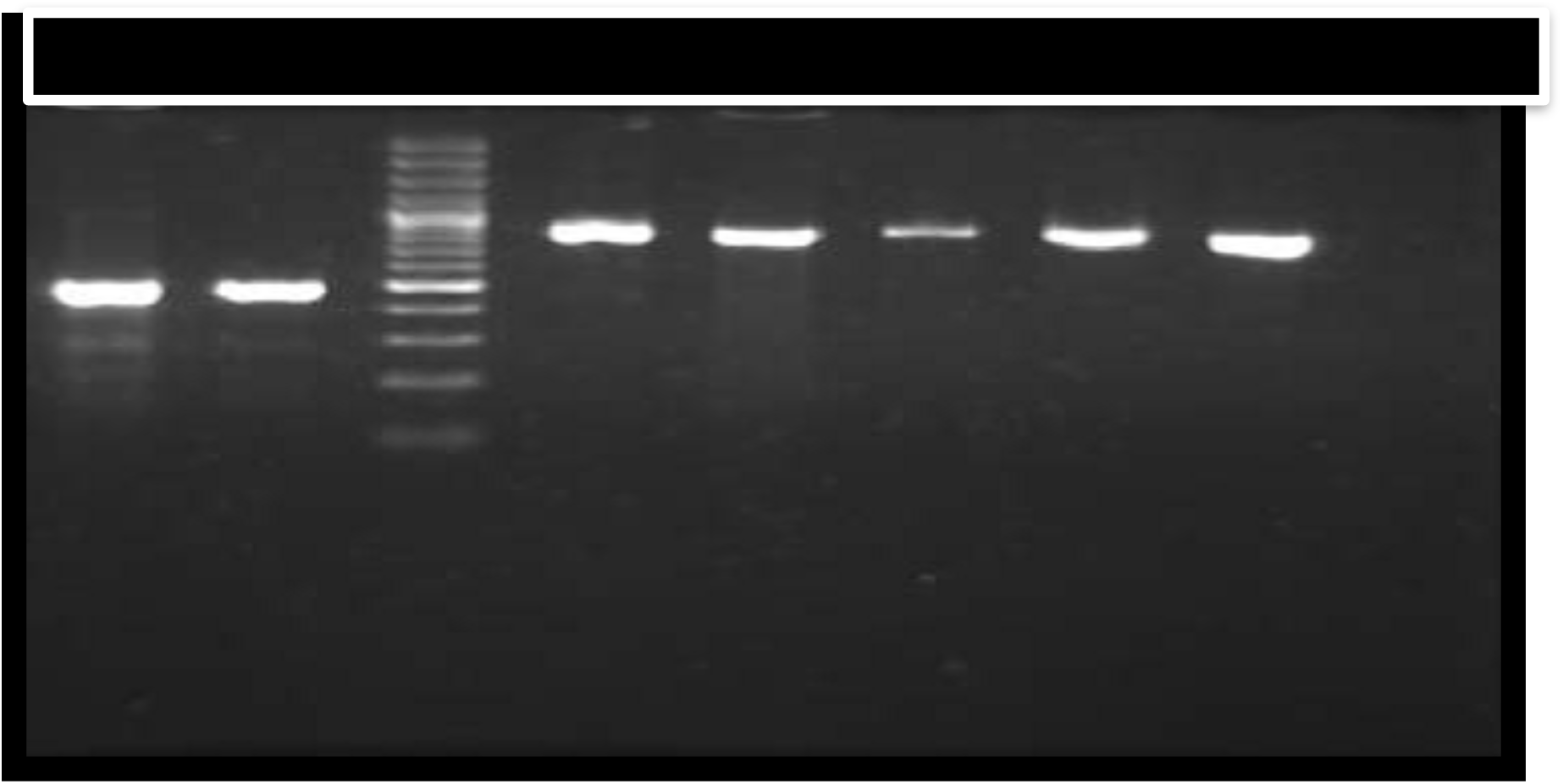
The PCR amplification of SHV and TEM genes: Lane number (1) showed positive SHV and Lane (2) is Control positive with a 471bp fragment of SHV gene .and Lane 3 is the 1500pb of DNA Ladder. Lane number 4 is Control positive showing a 861bp fragment of TEM gene. Lane numbers 5, 6, 7 and 8 show positive TEM gene.

Overall, ESBL producing strains were found in 85% (85/100) of various clinical specimens. The highest was in blood 100% (6/6) as well as stool 100% (4/4) followed by sputum 91% (21/23), wound 85% (12/14) and urine 82%(40\49) samples, while the lowest was noted in CSF 50% (2/4) [Table 2]. TEM gene was detected in 9.4% (8/85) of ESBL positive isolates. While SHV gene was found in 1.2% (1/85) of ESBL producing strains and in 6.7% (1/15) of ESBL-negative strains [Table 3].

**Table 2:**
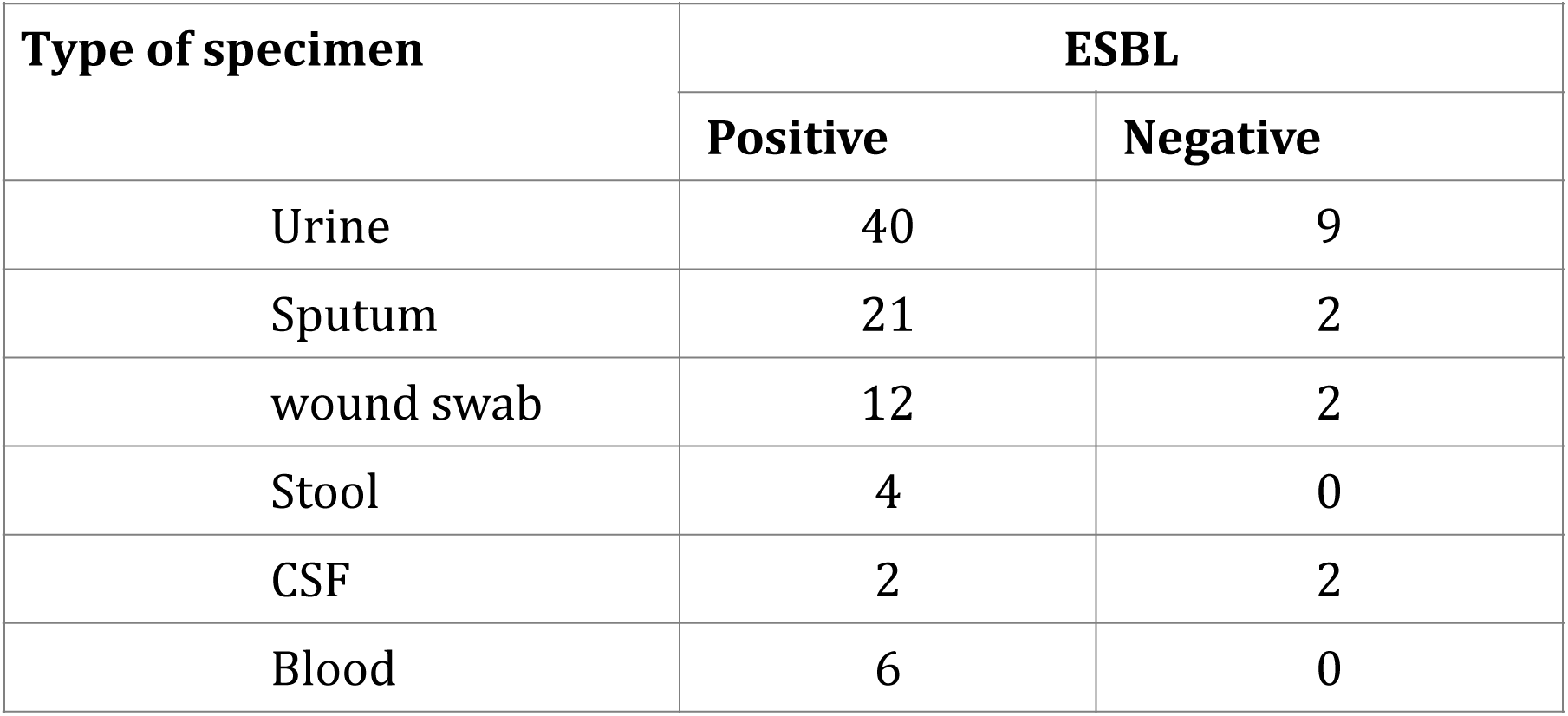
Distribution of ESBL among different specimens.

| Type of specimen | ESBL |  |
| --- | --- | --- |
|  | Positive | Negative |
| Urine | 40 | 9 |
| Sputum | 21 | 2 |
| wound swab | 12 | 2 |
| Stool | 4 | 0 |
| CSF | 2 | 2 |
| Blood | 6 | 0 |

**Table 3:** the distribution of SHV and TEM among both ESBL and non-ESBL.

| Targeted genes |  | ESBL |  |
| --- | --- | --- | --- |
|  |  | Positive | Negative |
| SHV | Positive | 1 | 1 |
|  | Negative | 0 | 0 |
| TEM | Positive | 8 | 0 |
|  | Negative | 77 | 0 |

## Discussion

Resistance to beta-lactam antibiotics of Gram negative bacteria isolated from clinical samples has been increased worldwide (22). The aim of this study was to investigate the prevalence of ESBL producing *K. pneumoniae* and distribution of ESBL-encoding genes, mainly TEM and SHV genes among clinical isolates. The present study showed that the prevalence of ESBL producing *K. pneumoniae* was 85%. This finding was higher than that all previously reported results in Sudan (59.6%) (17), India (44.4%) (23) and Taiwan 29.8% (24), Thailand (64%) (25), Korea (22.5) (26), Pakistan (36% (27) and Iran 52% (28). We suggest that the difference in the prevalence of ESBL is due to the un-uniformity of specimens when comparing with previous studies.

From a molecular prospective view our study marked a superiority of TEM gene to SHV (9.4% to 1.2%, respectively) in ESBL positive isolates. This was similar finding documented in Iran where TEM gene to SHV gene was (46.4% to 11.2%, respectively) (29). Unlike, a number of studies conducted in different countries, Namely, Sudan 17.6% SHV and 16.2% for TEM (17)., Saudi Arabia; 89.1% SHV and 70.9% TEM (30) and Turkey; 74.3% SHV and 52.7% TEM, (31). Interestingly, SHV gene was also detected in 6.7% of non-ESBL producing *K. pneumoniae.* This finding was not novel because it was reported in studies carried out in Saudi Arabia (30) and Switzerland (32). However, the variation in findings is thought to be due to; other mechanisms by which resistance evolves, other genes encoding for ESBL-production or mutations in ESBL-genes, such as carrying the gene encoding for ESBL-production but lacking the ability to translate it in order to give the functioning phenotype.. Furthermore, our study also detected a high prevalence of Meropenem resistance (33%) which is by far higher than the figures reported in a study in Canadian hospitals 24.1% (33). Besides, a substantial resistance was observed to Colistin (88%) almost similar to a study conducted in Italy where 58 colistin-resistant *K. pneumoniae* isolates were recovered (34).

## Conclusion

This study concludes that the prevalence of ESBLs producing *K. pneumoniae* was high in various clinical specimens. TEM and SHV genes were existed in certain ESBLs producing *K. pneumoniae.* Therefore, in-depth molecular studies are required to understand the resistance mechanisms of ESBLs producing *K. pneumoniae*.

## Data Availability

Contact the author

## Ethics

This study was approved by the Medical Specialization Ethics Review Board, Sudan.

